# Quantifying movement reserve in multiple sclerosis via diurnal activity quantiles

**DOI:** 10.1101/2025.05.05.25327033

**Authors:** Pratim Guha Niyogi, Muraleetharan Sanjayan, Rahul Ghosal, Jeff Goldsmith, Kate Fitzgerald, Ellen Mowry, Vadim Zipunnikov

**Author notes:** Ellen Mowry and Vadim Zipunnikov contributed equally as co-senior authors.

## Abstract

**Background:** Conventional clinical assessments in multiple sclerosis (MS), such as the Expanded Disability Status Scale (EDSS), often miss subtle functional changes. While accelerometry provides an objective measure of real-world motor activity, most daily summaries focus on average values, neglecting both peak performance and its variability throughout the day. This diurnal peak performance variability may reflect a person’s capacity to sustain high-effort activity despite fatigue, a phenomenon we term observable movement reserve.

**Objective:** To evaluate whether upper diurnal activity quantiles derived from accelerometry data quantify observable movement reserve, and to examine their association with EDSS-measured disability in MS.

**Methods:** In a cohort of 248 adults with MS (mean age 54.8 years, 71% female; EDSS range 0–6.5), continuous wrist accelerometry was collected over two weeks. We used novel scalar-on-function regression (SOFR) to compare several diurnal activity characteristics: mean, variability, and 50th–100th percentiles. SOFR models adjusted for age, sex, and BMI, and their cross-validated *R*^2^ were used to assess the strength of the association between EDSS and each diurnal activity curve.

**Results:** The upper diurnal activity quantiles, particularly the 95th to 100th percentiles, demonstrated the strongest association with EDSS, outperforming both diurnal mean and variability-based diurnal activity curves (crossvalidated *R*^2^ increased from 0.12 for the diurnal mean to 0.22 for the diurnal 100th percentile). The largest contribution to predictive power came from the late afternoon and evening hours, highlighting the importance of time-of-day in assessing disability.

**Conclusion:** Diurnal peak activity provides a sensitive, time-of-day–specific measure of observable movement reserve that closely aligns with EDSS-measured disability. This reserve fluctuates across the day, likely reflecting circadian patterns of energy and fatigue. By capturing both the timing and intensity of peak activity, the proposed metrics offer a clinically meaningful tool for monitoring functional change over time and may enhance the ability to track disease progression in MS.

## 1 Introduction

Multiple sclerosis (MS) is a chronic autoimmune disease of the central nervous system that leads to demyelination and neurodegeneration, resulting in symptoms such as mobility issues, fatigue, and cognitive impairment (Confavreux and Compston, 2009). Disability in MS is commonly assessed using the Expanded Disability Status Scale (EDSS), a standardized clinical scale ranging from 0 (normal neurological exam) to 10 (death due to MS) in 0.5-point increments While EDSS provides a snapshot of overall disability, it may overlook subtle, real-world fluctuations in functional capacity, particularly fatigue-related within-day declines that often occur later in the day and may go unrecorded in clinical exams. These fluctuations may reflect a person’s capacity to generate and sustain effortful movement despite fatigue, a concept we refer to as observable movement reserve. Capturing this reserve in everyday settings may provide a more sensitive and dynamic assessment of disability.

Clinicians currently lack practical tools to measure this reserve in real-world settings. Capturing it through wearable-based activity metrics may enable more sensitive detection of fluctuations in functional decline. Wrist-worn actigraphs provide continuous monitoring of physical activity in free-living conditions over extended periods of time (Ancoli-Israel et al., 2003; Casey et al., 2018; Sparaco et al., 2018; Evans et al., 2021). Despite their potential, most analytic approaches reduce activity to daily averages or time-above-threshold summaries. By failing to capture both the full range of activity intensities and their diurnal variation, these approaches obscure critical information about the magnitude and timing of activity, key features that are often linked to fatigue and energy fluctuations as well as behavioral rhythms in MS. As a result, current approaches may fall short in fully leveraging the richness of multi-day accelerometry data to assess everyday activity reserve, and its relationship with disability progression in MS.

We hypothesize that modeling time-specific upper activity levels can provide a window into observable movement reserve, an individual’s capacity to produce high-effort activity in daily life, particularly during traditionally higher-fatigue periods such as late afternoon and evening. To explore the question of when during the day people with MS are most functionally limited, diurnal activity patterns can be analyzed using functional data analysis (FDA) (Wang et al., 2016; Crainiceanu et al., 2024). Prior MS studies using FDA have shown that evening activity patterns are closely linked to disability (Keller et al., 2022; Bou Rjeily et al., 2025; Fitzgerald et al., 2025). However, these approaches typically rely on mean curves and overlook variation in intensity, which may obscure meaningful within-day changes.

Time-varying quantile functions, recently introduced by (Ghosal et al., 2022; Méndez-Civieta et al., 2025), extend traditional FDA by capturing the full range of activity intensities at a specific time of the day. Unlike mean-based approaches, they characterize both typical and maximal/extreme activity levels. This fuller representation may more accurately quantify observable movement reserve, particularly in MS, where fatigue and function vary throughout the day.

Figures 1–3 compare two individuals with MS: one with low disability (EDSS = 1) and one with higher disability (EDSS = 6). While median activity levels (q = 0.5) differ between them, upper diurnal quantiles (90th–100th percentiles) reveal clearer distinctions, especially in the late afternoon and evening. The less disabled individual shows sustained late-day peaks, whereas the more disabled profile flattens earlier. Thus, upper quantiles may help to uncover time-specific reductions in movement reserve less visible in traditional summaries.

**Figure 1.**
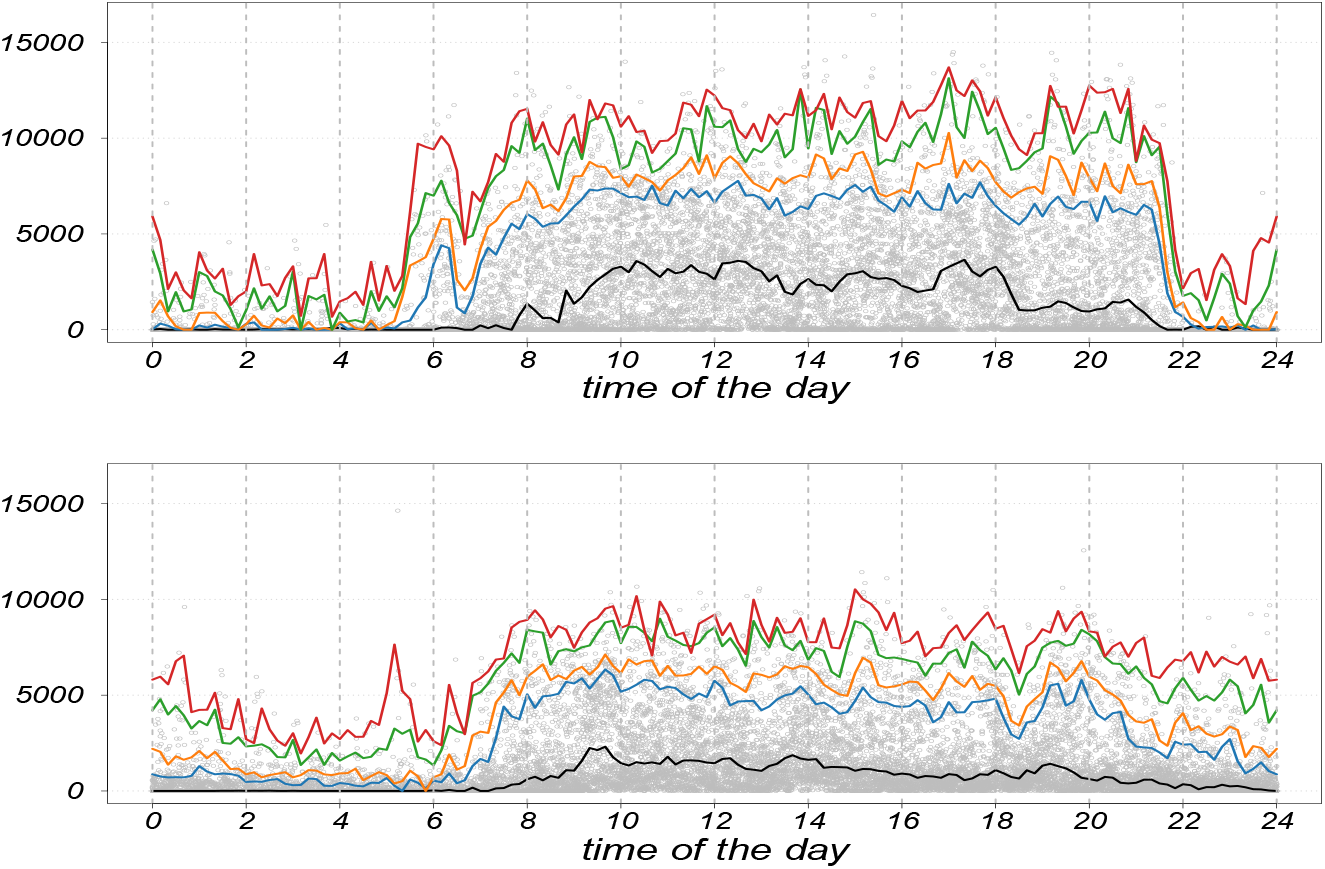
Quantile profile (in original scale) representing the distribution of two female subjects with EDSS scores 1 (low disability) and 6 (higher disability), respectively, for (*q* = 0.50, 0.90, 0.95, 0.99, 1).

Observable movement reserve refers to an individual’s capacity to generate and sustain peak activity throughout the day. We hypothesize that diurnal peak activity, captured using time-varying quantiles, provides a physiologically relevant and interpretable measure of this reserve. Using scalar-on-function regression, we assess how these diurnal activity quantiles relate to EDSS and evaluate their potential as clinically useful markers of functional variation in MS.

## 2 Methods

### 2.1 HEAL-MS study

We analyzed baseline data from the Home-based Evaluation of Actigraphy to Predict Longitudinal Function in MS (HEAL-MS) observational cohort, recruited at Johns Hopkins University between January 2021 and March 2023 (Tian et al., 2020; Bou Rjeily et al., 2025). The study enrolled patients from the Johns Hopkins MS Precision Medicine Center of Excellence. The analysis included 248 participants, all aged at least 40 (mean age 54.8 years, SD 8.5, 70.7% are female), with no known comorbidities affecting physical activity such that changes in the accelerometry measures were more likely to reflect the impact of MS itself, rather than being influenced by other comorbid conditions. The eligibility criteria further required participants to have MS, with no relapse occurring within the six months prior to enrollment, so that recover from a recent relapse did not affect the baseline measurements, and baseline EDSS scores at most 6.5, corresponding to the ability to, at minimum, walk with a walker. A third of the cohort was selected because they fulfilled criteria for progressive MS, but the majority was still classified as having relapsing-remitting MS, though they were at an age where the transition to progressive MS might occur within a few years.

### 2.2 Accelerometry data

Participants wore a GT9X ActiGraph device on the non-dominant wrist continuously for two weeks. This triaxial accelerometer recorded raw acceleration data at a 30 Hz sampling rate, which was then aggregated into 1-minute activity count (AC) epochs using ActiLife v6.13.4 (Lite edition). The resulting .gt3x files were processed using the `read.gt3x` package in R, yielding daily physical activity profiles in a 1440-minute analytic format.

Data quality was ensured through standard preprocessing including the following steps. Non-wear time was defined as any continuous 90-minute interval of zero counts. A valid day was defined as having at least 90% wear time, and each participant was required to have at least three valid days of data.

Daylight Saving Time (DST) adjustments were also implemented. For spring shifts (1-hour loss), the missing hour was imputed by averaging the same hour across other days for that participant. For fall shifts (1-hour gain), duplicate values were averaged. Any missing hour on the final day was also imputed using the same-hour average from other days.

In addition to the original AC scale, the data were log-transformed using the function *x →* log(*x* + 1) to reduce skewness and stabilize variance.

### 2.3 Expanded Disability Status Scale

The EDSS examination was administered by a physician trained in EDSS assessment who remained blinded. The modified MS Functional Composite (MSFC), incorporating the 9-hole peg test, timed 25-foot walk, high- and low-contrast letter acuity (using binocular 2.5% contrast Sloan charts), and the Symbol Digit Modalities Test (SDMT), was conducted by a trained, blinded study coordinator. Additionally, participants completed patient-reported outcomes, including the Patient-Determined Disease Steps (PDDS), Neuro-QoL, MS Impact Score-29 (MSIS-29), and the International Physical Activity Questionnaire (IPAQ). See Bou Rjeily et al. (2025) for more details.

### 2.4 Diurnal activity curves

Time-varying characteristics of accelerometry data can be summarized using a range of aggregation methods that capture different aspects of daily activity patterns. Common approaches include calculating the mean to estimate central tendency and, more recently, the variance to quantify fluctuation. In our approach, we propose to additionally calculate quantiles to quantify the distribution of activity intensity. Particularly, upper quantiles may be better tracking peak performance and activity reserve. These summary measures can be computed within defined time windows (10-minute, hour). In the context of MS, such temporal features may offer a novel way to quantify real-world activity.

We focus on 10 min time bins and calculate diurnal curves using standard temporally local distributional summaries. Suppose *y*_*ijst*_ is the minute-level activity count (AC) or log-transformed activity count (LAC) of the *i*-th subject, *j*-th day, *s*-th bin, and *t*-th time-point within the bin. Thus, consider *y*_*ijst*_ = *µ*_*is*_ + *σ*_*is*_*ϵ*_*ijst*_ where *µ*_*is*_ the bin-specific mean and the centered random noise *ϵ*_*ijst*_ with variance 1, where *σ*_*is*_ is the bin-specific standard deviation. For each temporal local bin, we calculate the various distributional summaries outlined below.

(A) subject-specific mean defined on the bin *s, µ*_*i*_(*s*); where 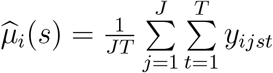.

(B) subject-specific mode defined on the bin *s, mode*_*i*_(*s*).

(C) subject-specific standard deviation defined on the bin *s, σ*_*i*_(*s*),

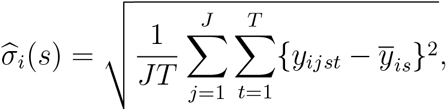

where 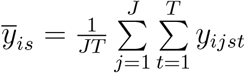.

(D) subject specific median absolute deviation on the bin *s, mad*_*i*_(*s*),

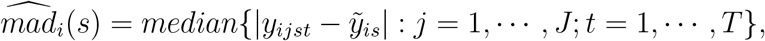

where 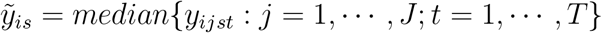. This is a robust measure of the viability.

(E) subject-specific coefficient of variation defined on the bin *s, 𝒞*_*i*_(*s*) = *σ*_*i*_(*s*)*/µ*_*i*_(*s*), where *µ*_*i*_(*s*) and *σ*_*i*_(*s*) are the subject-specific mean and standard deviation at bin *s*. For the purpose of computation of the coefficient of variation, if either *µ*_*i*_(*s*) or *σ*_*i*_(*s*) is zero for a given bin *s* and the threshold *c*, we define that *𝒞*_*i*_(*s*) = 0. This measures the relative dispersion of the data around the mean. A low coefficient of variation indicates that activity levels are relatively consistent across time-points for fixed time bins and therefore, this indicates that the subject tends to have a similar level of physical activity consistently in that time bins. A high coefficient of variation suggests greater variability in activity levels across time-points in each time-bins.

(F) subject-specific quantiles of AC (or LAC) defined on the bin *s, q*_*i*,*k*_(*s*); the *k*-th percentile of AC (or LAC) ignoring the level of day and time within the bin *s*. In particular, for *k* = 100, we obtain the subject-specific maximum of the AC (or LAC) at bin *s*. This calculates the highest value (or peak level) of the physical activity across different time-points in a certain bin. In individuals with MS, this may vary widely depending on the severity of their symptoms, their level of disability, and their overall functional capacity. A higher maximum count may indicate better physical function and endurance, while a lower maximum count may suggest limitations in mobility and activity. Besides the maximum, we consider *k* = 50, the median of AC (or LAC); *k ∈ {*90, 95, 99*}* for the 90, 95, and 99th quantile of AC (or LAC) for each bin.

Formally, we observe the data {*𝒟*_*i*_ = {EDSS_*i*_, *ℛ*_*i*_(*s*), age*i*, sex_*i*_, BMI_*i*_*}* : *s ∈* [0, 24]; *i* = 1, …, *n}*, where *ℛ*_*i*_(*s*) is the set of all subject-specific temporally local distributional characteristics such as mean, SD, MAD, coefficient of variation, quantiles of physical activity data. Furthermore, we assume that *𝒟*_*i*_s are independent and each of the components in *𝒟*_*i*_s are observed at every bin.

Figures 2 and 3, previously introduced as illustrative examples, show that both upper quantiles and traditional distributional summaries differentiate between individuals with low and high EDSS. The low-EDSS participant exhibits sustained peak activity later in the day, while the high-EDSS profile shows earlier decline and reduced variability in late afternoon and evening hours, consistent with reduced movement reserve.

**Figure 2.**
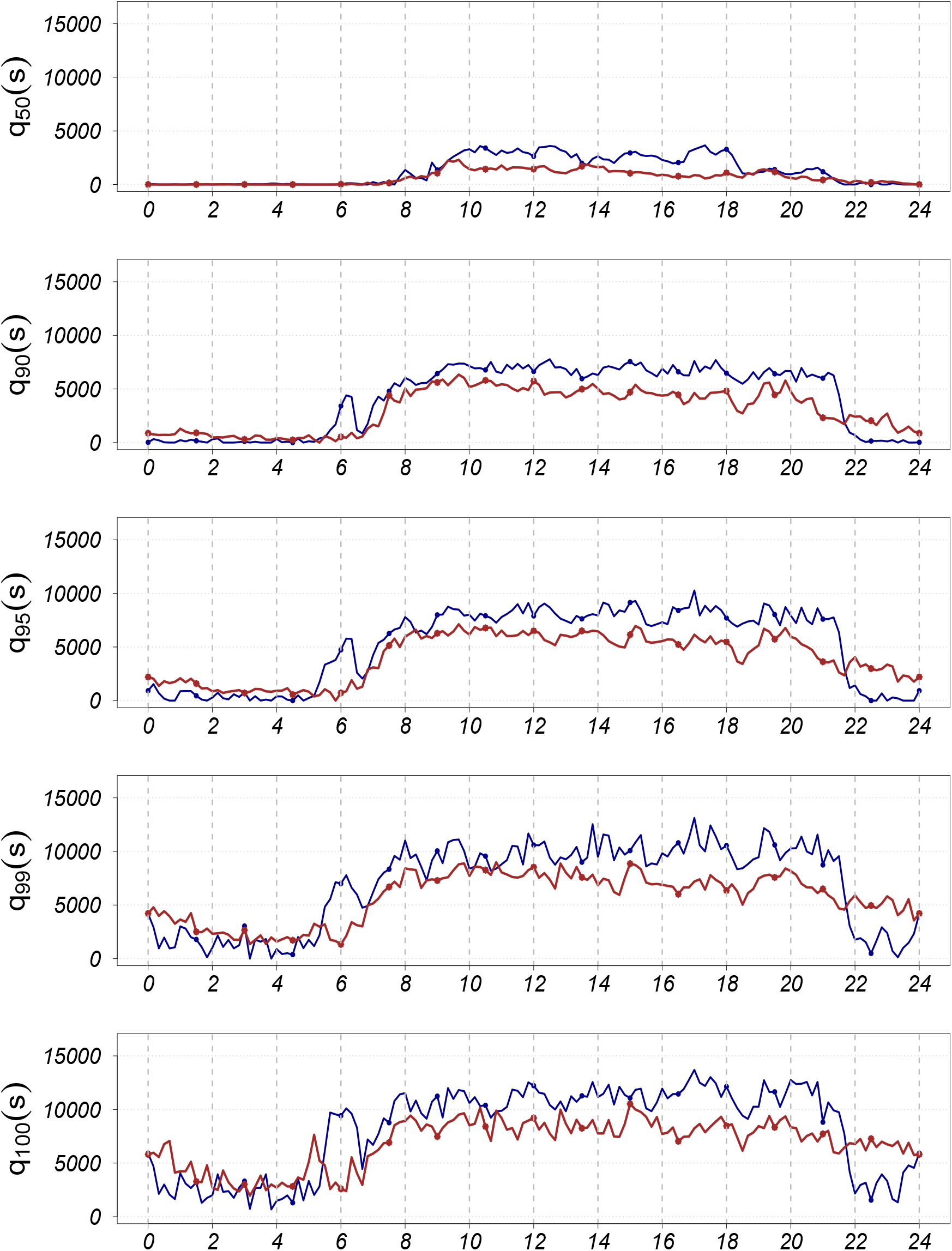
Comparison plots for quantile profiles (in original scale) of two subjects with EDSS scores 1 (low disability, color coded as blue) and 6 (higher disability, color coded as red), respectively, for (*q* = 0.50, 0.90, 0.95, 0.99, 1).

**Figure 3.**
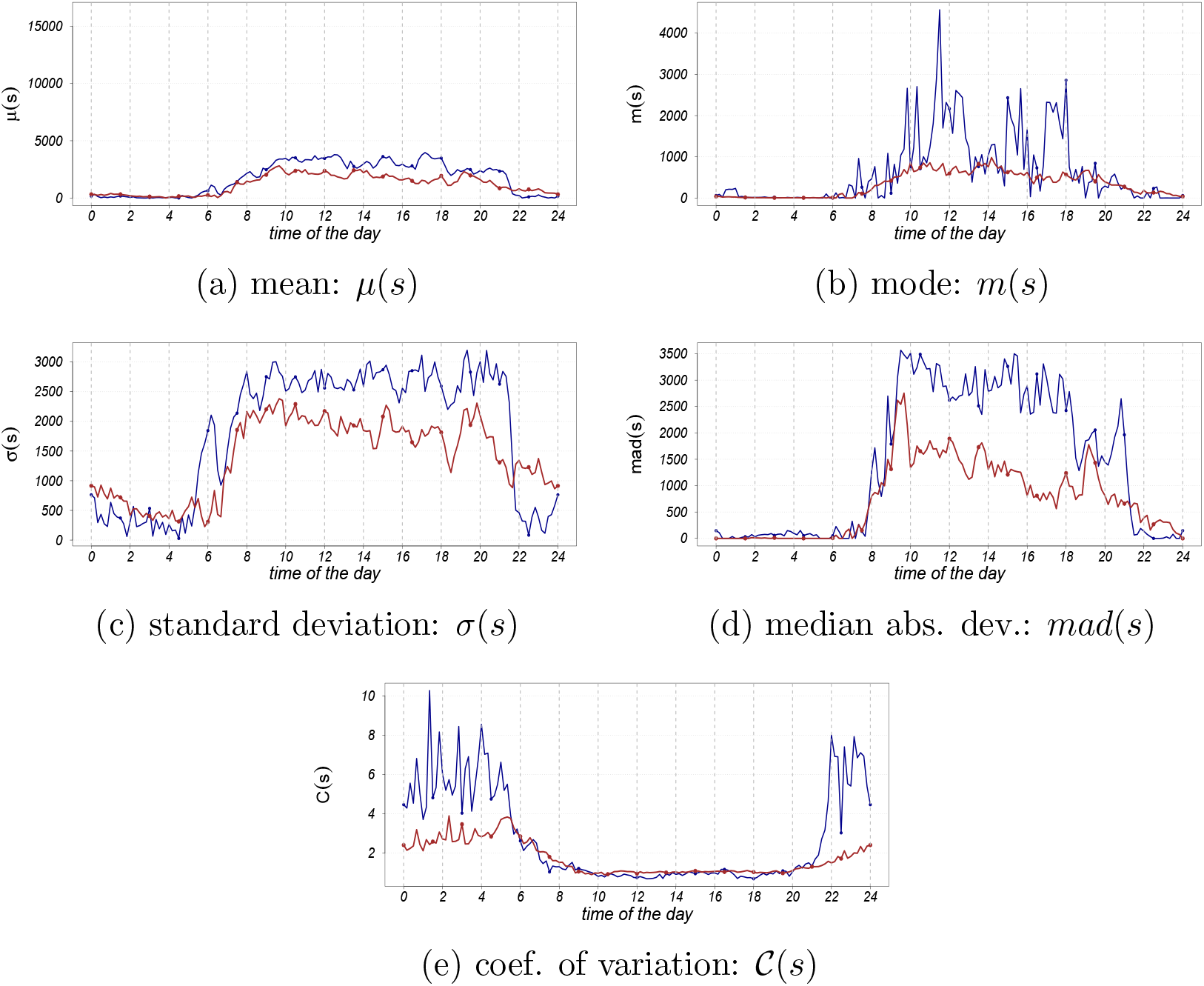
Mean, mode, standard deviation, median absolute deviation, coefficient of variations based on the activity counts (in original scale) for two subjects with EDSS score 1 (low disability, color coded as blue) and EDSS score 6 (higher disability, color coded as red).

Next, we formally assess the discriminative power of these distributional diurnal summaries across all study participants by modeling their association with EDSS using scalar-on-function regression.

### 2.5 Scalar-on-function regression

Scalar-on-function regression (SOFR) builds (Crainiceanu et al., 2024) a relationship between a scalar outcome (EDSS) and a time-varying function (participant-specific curve capturing distributional properties specific for a fixed time of the day). This allows us to assess how activity in the specific time of the day relate to disability. The SOFR models will be adjusted for age, gender, and BMI.

The scalar-on-function regression is a flexible, non-parametric data-driven technique that assesses the association between the subject-specific clinical characteristics (EDSS) and subject-specific diurnal activity summary. The SOFR model is mathematically described as follows:

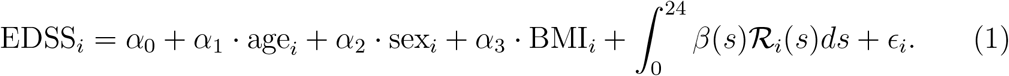

The model 1 estimates the adjusted relationship between the scalar outcome of interest (EDSS) and *ℛ*_*i*_(*s*), a diurnal activity curve. From a clinical perspective, scalar-on-function regression allows us to examine how activity patterns (quantified via time-varyng mean, variability, and quantile levels) relate to disability. Unlike traditional regression models that use one summary number, SOFR evaluates the entire daily activity curve, revealing when physical capacity diverges across patients with different EDSS scores.

We apply `pfr` function in R available in `‘refund’` package (Goldsmith et al., 2020). To ensure the reliability of our assessment, we employ a rigorous 5-fold cross-validation technique with 100 random replications and present the average cross-validated *R*^2^ along with a 95% confidence interval.

## 3 Results

We evaluated how well time-of-day–specific diurnal activity curves with different temporallly local distributional aggregation predict EDSS scores using scalar-on-function regression, adjusting for age, sex, and BMI. To assess predictive performance, we applied 5-fold cross-validation (repeated 100 times) and report mean *R*^2^ values and 95% confidence intervals in Table 1.

**Table 1:** Cross-validated *R*^2^ obtained from scalar-on-function regression adjusted by age, gender, and BMI for predicting EDSS scores with local distributional characteristic metrics of physical activity in the original and log scales.

| characteristic metric | cross-validated $R^2$ | |
| --- | --- | --- |
|  | original scale | log scale |
| mean, $\mu(s)$ | 0.12 (0.09, 0.15) | 0.09 (0.06, 0.12) |
| mode, $m(s)$ | 0.05 (0.03, 0.08) | 0.10 (0.07, 0.13) |
| standard deviation, $\sigma(s)$ | **0.21 (0.18, 0.25)** | 0.04 (0.01, 0.06) |
| median absolute deviation, $mad(s)$ | 0.09 (0.07, 0.13) | 0.03 (0.01, 0.07) |
| coefficient of variation, $\mathcal{C}(s)$ | 0.05 (0.03, 0.08) | 0.06 (0.03, 0.09) |
| 50th quantile, $q_{50}(s)$ | 0.06 (0.04, 0.09) | 0.10 (0.07, 0.12) |
| 90th quantile, $q_{90}(s)$ | **0.19 (0.15, 0.23)** | **0.17 (0.13, 0.19)** |
| 95th quantile, $q_{95}(s)$ | **0.20 (0.16, 0.23)** | **0.18 (0.15, 0.21)** |
| 99th quantile, $q_{99}(s)$ | **0.21 (0.18, 0.24)** | **0.22 (0.19, 0.25)** |
| maximum, $q_{100}(s)$ | **0.22 (0.19, 0.25)** | **0.24 (0.21, 0.27)** |

Across both raw AC and LAC, upper quantiles consistently outperformed traditional metrics. For example, on the AC scale, the cross-validated *R*^2^ increased from 0.12 for the mean and 0.05 for the mode to 0.22 for the 100th percentile. The 90th to 99th percentiles also performed well, with *R*^2^ values between 0.19 and 0.21. Results were similar on the LAC scale, where the 100th percentile achieved an *R*^2^ of 0.24, more than doubling that of the mean (0.09). These findings suggest that capturing the highest levels of activity, rather than daily averages, provides more sensitive markers of disability in MS. Notably, standard deviation on the AC scale also performed well (*R*^2^ = 0.21), but its predictive power dropped on the LAC scale (*R*^2^ = 0.04), indicating sensitivity to transformation and limited robustness. In contrast, upper quantiles remained stable across both scales, highlighting their utility as transformation-invariant markers of functional capacity. Overall, traditional diurnal summaries explained only 9–11% of the variance in EDSS scores, whereas upper quantiles explained up to 24%, supporting their use as interpretable and robust indicators of observable movement reserve in MS.

Figure 4 illustrates the coefficient function for the 100th percentile. The strongest negative associations occur in the late afternoon and evening, indicating that individuals able to sustain high activity during these hours tend to have lower disability. This further supports the role of late-day peak activity as a marker of observable movement reserve. It also shows higher nighttime activity (approximately 12:00AM–4:00AM) was positively associated with EDSS scores. This suggests that individuals with greater disability may exhibit elevated nocturnal movement, potentially reflecting sleep disturbances or disrupted rest–activity rhythms. Time-of-day specific effects for all metrics are visualized in Figures S.1–S.7, highlighting where and when diurnal features relate most strongly to EDSS. Red regions indicate significant negative associations; blue regions indicate positive ones. To enhance interpretability, we also derived scalar biomarkers from the SOFR framework as 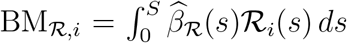, where *ℛ*_*i*_(*s*) is any local distributional metric described in Section 2.4. Figures S.4 and S.8 show the distribution of these biomarkers stratified by EDSS thresholds.

**Figure 4.**
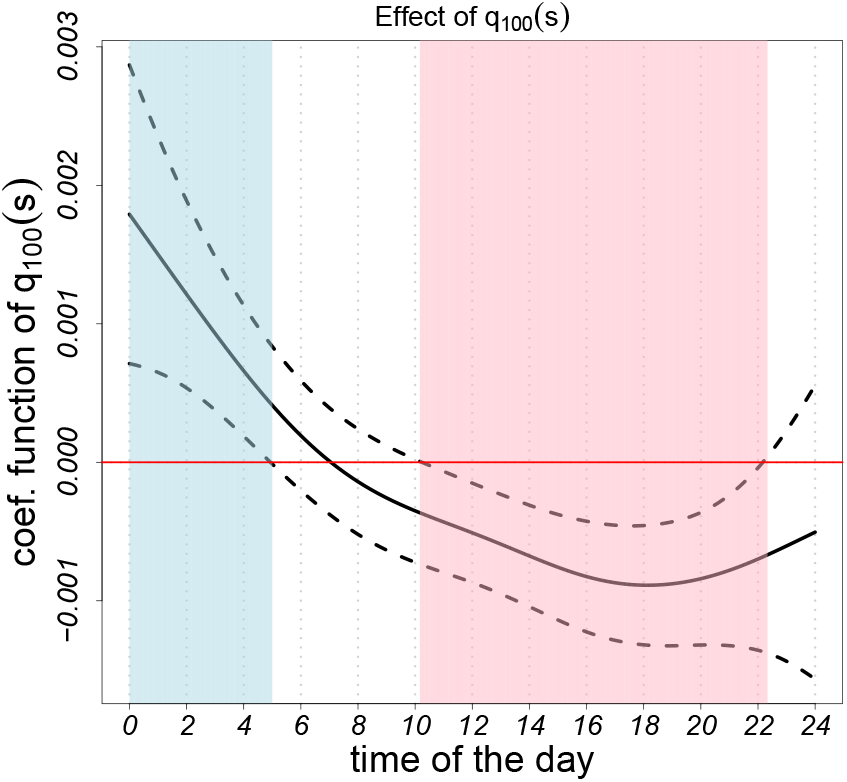
The estimated regression coefficient for models employing temporally local maximum based on activity counts (in original scale) is derived through scalar-on-function regressions to analyze EDSS scores. These functions are generated using activity counts during periods of activity. The solid black curve represents the estimated coefficient function, while dotted black lines denote the 95% confidence intervals.

Supplementary Tables S.1 and S.2 summarize the significance of model coefficients across metrics. The coefficients for the upper quantiles, especially the 90th, 95th, 99th, and 100th percentiles, were highly significant (*p <* 0.001), while age was also consistently associated with EDSS; sex and BMI did not show a consistent effect.

In summary, our findings suggest that patients who are able to sustain higher-intensity activity during late-day hours tend to have lower disability, highlighting the clinical relevance of observable movement reserve. The biomarkers derived from scalar-on-function regression provide interpretable, individualized measures of this reserve and diurnal motor capacity. Importantly, they could serve as digital indicators of fatigue susceptibility or functional resilience, enabling clinicians to track meaningful within-person changes over time, potentially identifying early signs of decline even when traditional measures like EDSS remain unchanged.

## 4 Discussion

This study presents a novel analytic framework for modeling accelerometry-derived diurnal patterns of peak activity in Multiple Sclerosis. By modeling participant-specific, time-varying activity curves and applying scalar-on-function regression (SOFR), we show that upper diurnal quantiles are more strongly and consistently associated with EDSS scores than traditional mean- or variability-based summaries.

A central contribution of this work is the introduction of observable movement reserve, the capacity to sustain high-effort activity across the day. We found that this reserve declines predictably in individuals with greater disability, particularly in the late afternoon and evening. The ability to maintain late-day activity may reflect resilience against fatigue and serve as a sensitive marker of early or subclinical decline, even when EDSS scores remain stable. These findings offer a more granular view of disability and complement prior studies that relied on mean-based diurnal activity curves, which are more limited in their ability to capture movement reserve (Keller et al., 2022; Bou Rjeily et al., 2025).

Our analysis leveraged time-varying quantile curves and functional regression models to capture the full intensity range and timing of daily activity. Upper quantiles showed a 2.5-fold improvement in cross-validated *R*^2^ over mean-based models, with the strongest associations localized to late-day hours. This supports the clinical relevance of peak activity as a marker of preserved functional reserve. Additionally, a positive association between nocturnal peak activity and EDSS suggests that disrupted nighttime movement patterns may serve as a complementary indicator of functional impairment in MS.

This study has several limitations that should be considered. First, the cross-sectional design restricts our ability to assess how observable movement reserve evolves over time or predicts future changes in disability. Second, our sample was limited to ambulatory individuals with EDSS scores of 6.5 or lower, and the findings may not generalize to individuals with more advanced disability or those using assistive devices. Third, while we adjusted for age, sex, and BMI, other potentially important factors such as comorbidities, pain, depression, and motivation may influence daily activity levels and were not explicitly accounted for. Lastly, although we interpret late-day reductions in activity as reflecting fatigue, this hypothesis should be tested directly in future work using validated fatigue measures and patient-reported outcomes.

Our use of upper diurnal activity quantiles parallels the application of the 95th percentile of stride velocity (Q95) in Duchenne Muscular Dystrophy (DMD), where Q95 has been shown to be more sensitive to functional decline than mean- or median-based measures. Importantly, Q95 is derived from continuous, real-world ankle-worn accelerometry data and has been qualified by the European Medicines Agency (EMA) as an acceptable secondary and exploratory primary endpoint for clinical trials in DMD (Servais et al., 2023), highlighting regulatory recognition of quantile-based digital biomarkers. In both DMD and MS, upper quantiles capture peak performance, offering a window into functional reserves expressed under real-world conditions. In our approach, tracking quantiles diurnally further improves discriminatory power by identifying and emphasizing time-of-day–specific reductions in movement reserve.

While this study focuses on MS, the concept of observable movement reserve has broader relevance. In neurodegenerative diseases such as Parkinson’s and Alzheimer’s, early functional decline may manifest through reduced peak activity long before clinical scales detect change. Quantile-based modeling of wearable data may thus offer a generalizable framework for monitoring functional capacity and stratifying risk across a wide range of chronic and progressive conditions.

In summary, this study has important clinical implications by introducing observable movement reserve, derived from wrist-worn accelerometry, as a potential complement to EDSS in detecting subtle or fluctuating disability, particularly in individuals with mild symptoms or early signs of progression. By extending monitoring into patients’ daily lives, this approach offers clinicians a tool to track real-world peak activity over time, support individualized care, and improve patient stratification in clinical trials.

## Data Availability

All data produced in the present study are available upon reasonable request to the authors after a signed data use agreement and de-identification.

## Abbreviations

AC: activity counts
EDSS: expanded disability status scale
LAC: log activity counts
MAD: median absolute deviation
MS: multiple sclerosis
PA: physical activity
SD: standard deviation
SOFR: scalar-on-function regression

## Acknowledgements

This research was partially supported by the grant R01NR018851 from the National Institute of Health (NIH).

## References

Ancoli-Israel, S., R. Cole, C. Alessi, M. Chambers, W. Moorcroft, and C. P. Pollak (2003). The role of actigraphy in the study of sleep and circadian rhythms. Sleep 26(3), 342–392.

Bou Rjeily, N., M. Sanjayan, P. Guha Niyogi, B. E. Dewey, A. Zambriczki Lee, C. Hulett, G. Dagher, C. Hu, R. D. Mazur, E. M. Kenney, et al. (2025). Accelerometry-assessed physical activity and circadian rhythm to detect clinical disability status in multiple sclerosis: Cross-sectional study. JMIR mHealth and uHealth 13, e57599.

Casey, B., S. Coote, and A. Donnelly (2018). Objective physical activity measurement in people with multiple sclerosis: a review of the literature. Disability and Rehabilitation: Assistive Technology 13(2), 124–131.

Confavreux, C. and A. Compston (2009). The natural history of multiple sclerosis. McAlpine’s multiple sclerosis, 183.

Crainiceanu, C. M., J. Goldsmith, A. Leroux, and E. Cui (2024). Functional data analysis with R. CRC Press.

Evans, M. A., D. J. Buysse, A. L. Marsland, A. G. Wright, J. Foust, L. W. Carroll, N. Kohli, R. Mehra, A. Jasper, S. Srinivasan, et al. (2021). Meta-analysis of age and actigraphy-assessed sleep characteristics across the lifespan. Sleep 44(9), zsab088.

Fitzgerald, K. C., M. Sanjayan, B. Dewey, P. Guha Niyogi, N. B. Rjeily, Y. Fadlallah, A. Delaney, A. Z. Lee, S. Duncan, C. Wyche, et al. (2025). Within-person changes in objectively measured activity levels as a predictor of brain atrophy in multiple sclerosis. medRxiv, 2025–01.

Ghosal, R., V. R. Varma, D. Volfson, J. Urbanek, J. M. Hausdorff, A. Watts, and V. Zipunnikov (2022). Scalar on time-by-distribution regression and its application for modelling associations between daily-living physical activity and cognitive functions in alzheimer’s disease. Scientific reports 12(1), 11558.

Goldsmith, J., F. Scheipl, L. Huang, J. Wrobel, C. Di, J. Gellar, J. Harezlak, M. W. McLean, B. Swihart, L. Xiao, C. Crainiceanu, and P. T. Reiss (2020). refund: Regression with Functional Data. R package version 0. 1-23.

Keller, J. L., F. Tian, K. C. Fitzgerald, L. Mische, J. Ritter, M. G. Costello, E. M. Mowry, V. Zippunikov, and K. M. Zackowski (2022). Using real-world accelerometry-derived diurnal patterns of physical activity to evaluate disability in multiple sclerosis. Journal of Rehabilitation and Assistive Technologies Engineering 9, 20556683211067362.

Mendez-Civieta, A., Y. Wei, K. M. Diaz, and J. Goldsmith (2025). Functional quantile principal component analysis. Biostatistics 26(1), kxae040.

Servais, L., D. Eggenspieler, M. Poleur, M. Grelet, F. Muntoni, P. Strijbos, and M. Annoussamy (2023). First regulatory qualification of a digital primary endpoint to measure treatment efficacy in dmd. nature medicine 29(10), 2391–2392.

Sparaco, M., L. Lavorgna, R. Conforti, G. Tedeschi, and S. Bonavita (2018). The role of wearable devices in multiple sclerosis. Multiple sclerosis international 2018(1), 7627643.

Tian, F., K. Fitzgerald, J. Keller, K. Zackowski, V. Zipunnikov, and E. Mowry (2020). A longitudinal study of objectively-measured physical activity patterns in multiple sclerosis. In MULTIPLE SCLEROSIS JOURNAL, Volume 26, pp. 25–25. SAGE PUBLICATIONS LTD 1 OLIVERS YARD, 55 CITY ROAD, LONDON EC1Y 1SP, ENGLAND.

Wang, J.-L., J.-M. Chiou, and H.-G. Muller (2016). Functional data analysis. Annual Review of Statistics and its application 3(1), 257–295.

