## Supplemental Figures and Tables for "Quantifying movement reserve in multiple sclerosis via diurnal activity quantiles"

### Supplementary material for ‘Quantifying movement reserve in multiple sclerosis via diurnal activity quantiles’

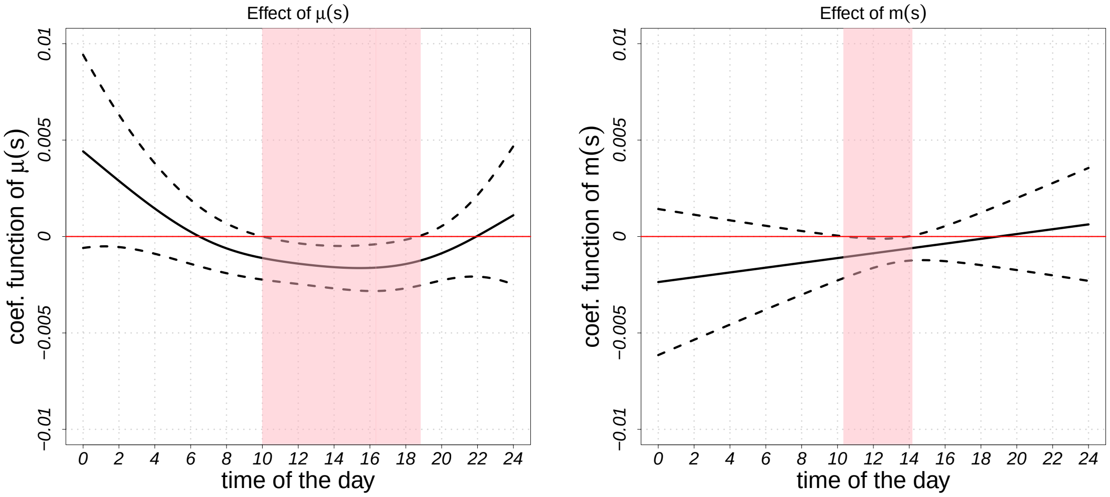


Figure S.1: The estimated regression coefficients for models employing temporally local summary functions such as mean, and mode based on activity counts (in original scale) are derived through scalar-on-function regressions to analyze EDSS scores. These functions are generated using log-activity counts during periods of activity. The solid black curve represents the estimated coefficient function, while dotted black lines denote the 95% confidence interval.

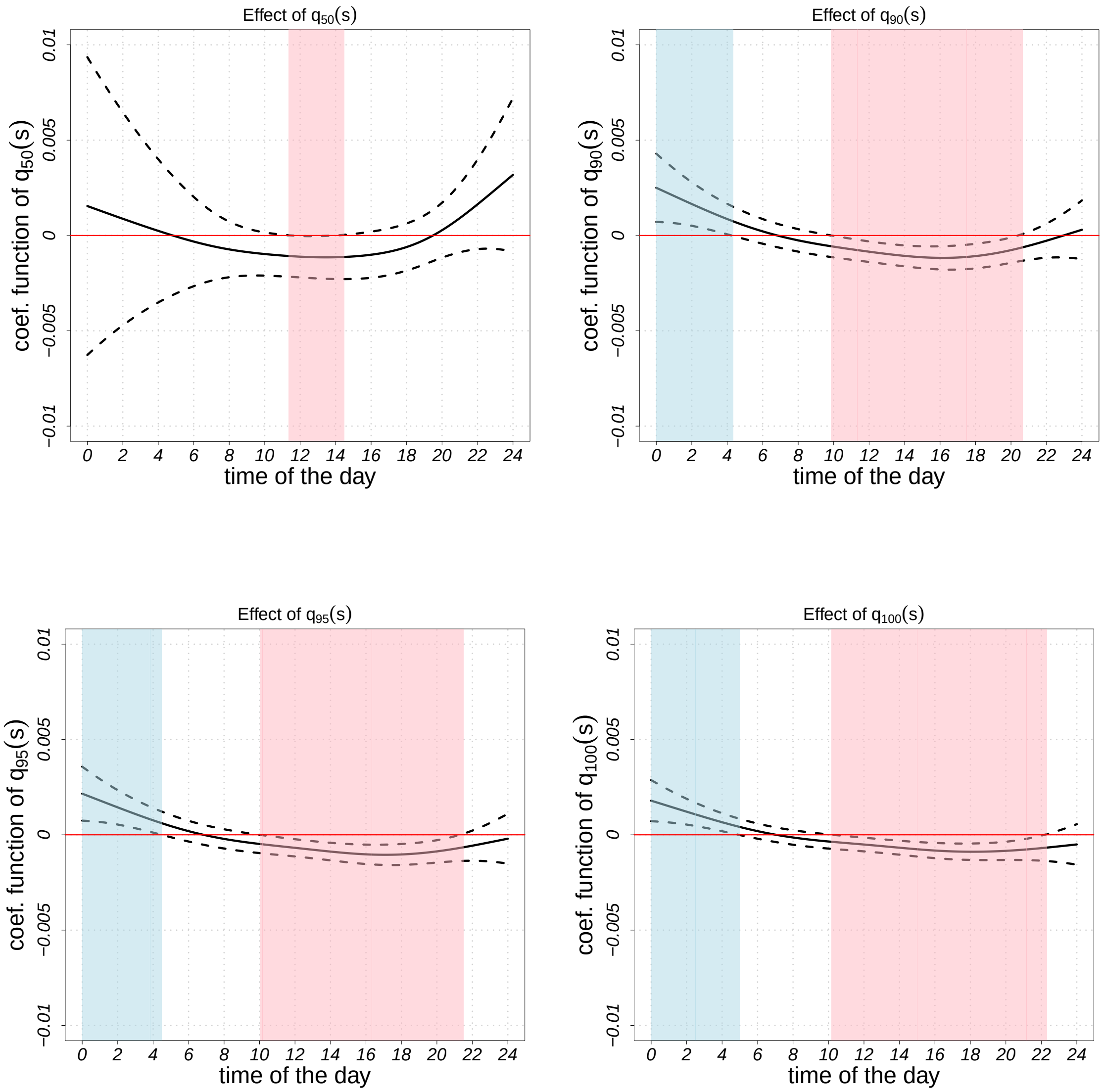


Figure S.2: The estimated regression coefficients for models employing temporally local quantile functions based on activity counts (in original scale) that are derived through scalar-on-function regressions to analyze EDSS scores. These functions are generated using log-activity counts during periods of activity. The solid black curve represents the estimated coefficient function, while dotted black lines denote the 95% confidence interval.

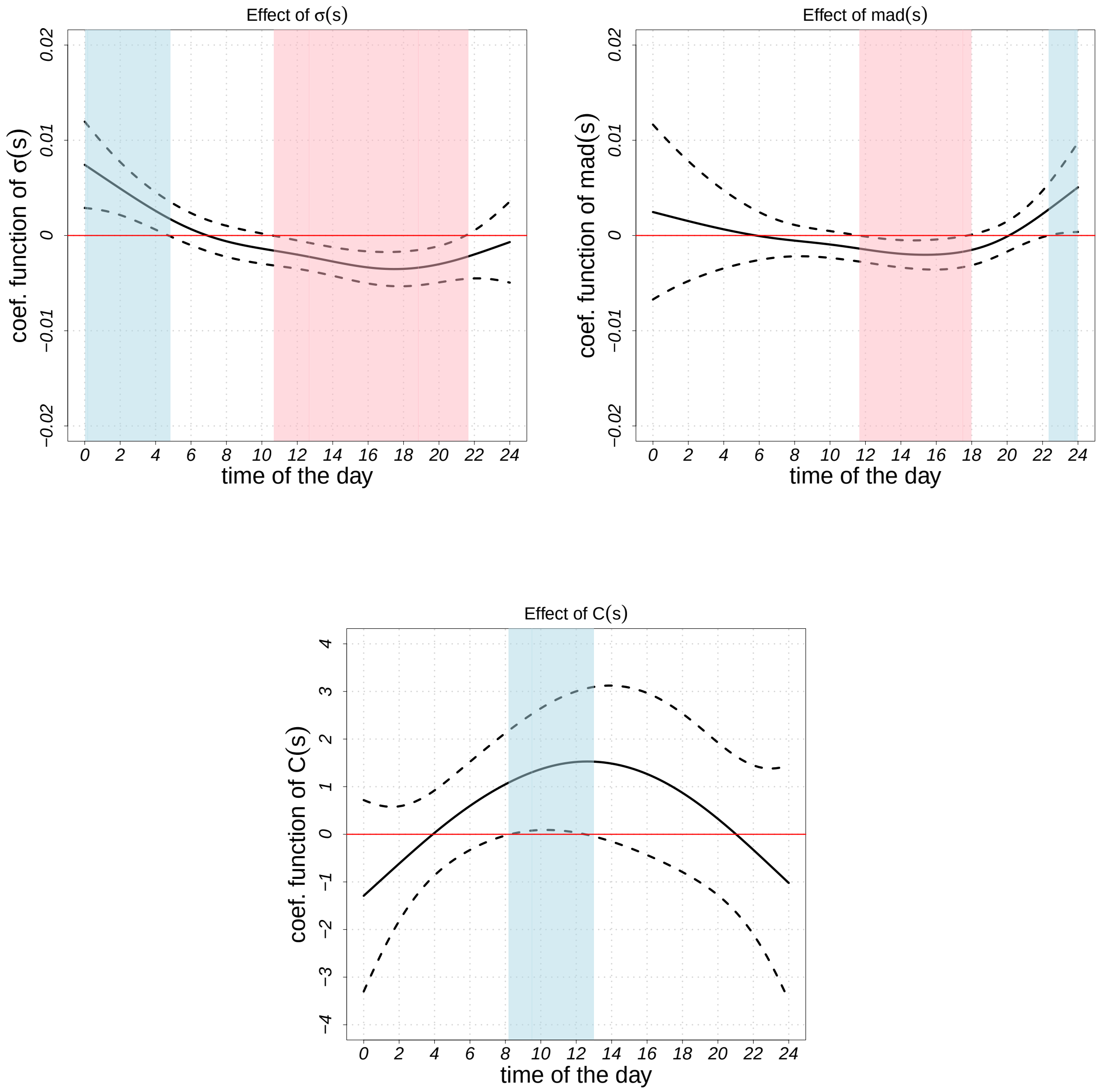


Figure S.3: The estimated regression coefficients for models employing temporally local variability functions such as standard deviation, median absolute deviation, and coefficient of variation based on activity counts (in original scale) are derived through scalar-on-function regressions to analyze EDSS scores. These functions are generated using log-activity counts during periods of activity. The solid black curve represents the estimated coefficient function, while dotted black lines denote the 95% confidence interval.

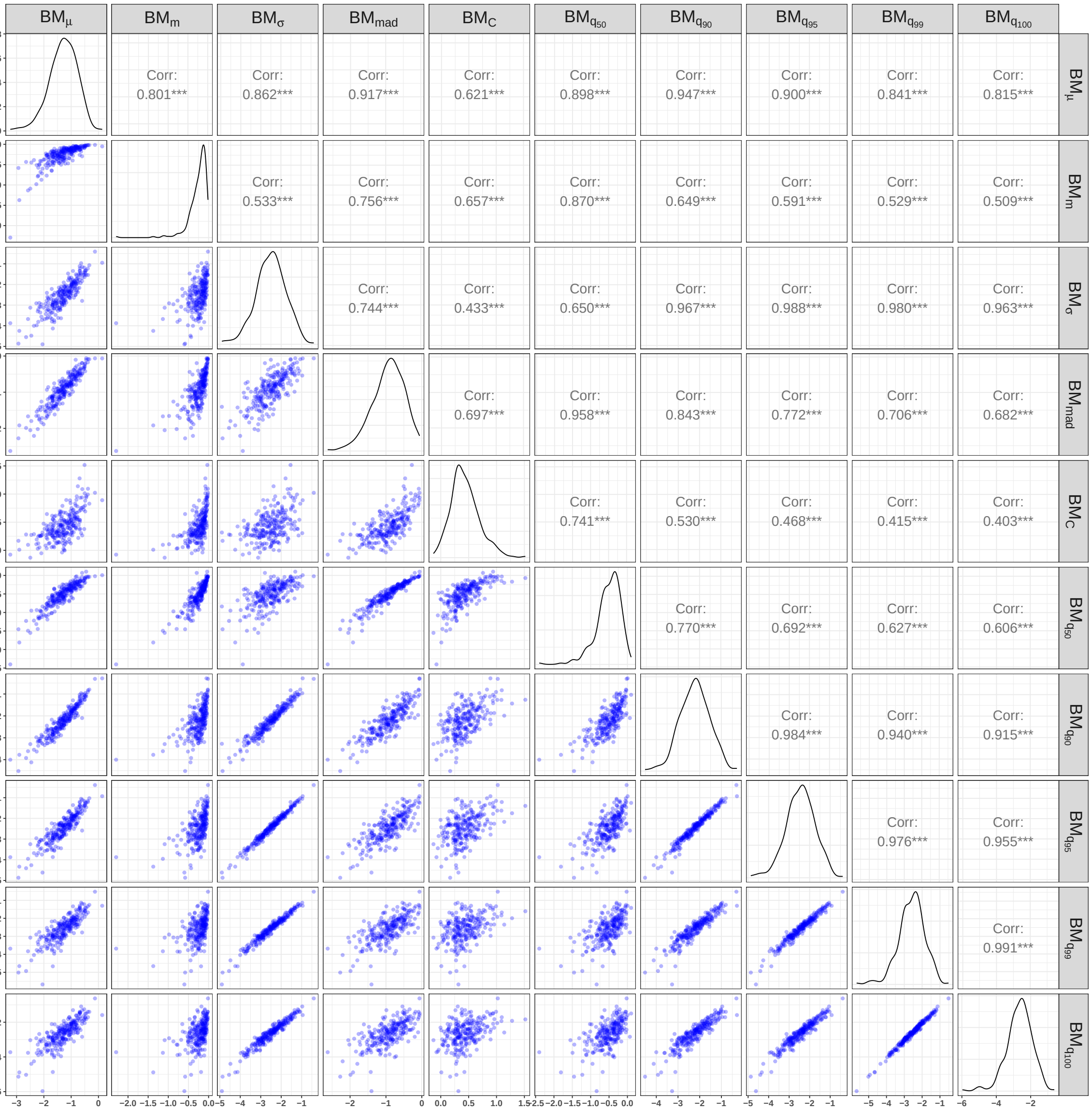


Figure S.4: Scatterplots of the estimated weighted scores corresponding to the predictor of different local distributional characteristic metrics, respectively. These local distributional characteristic metrics were calculated based on the physical activity counts in the original scale. All correlation coefficients indicate Spearman's rank correlation.

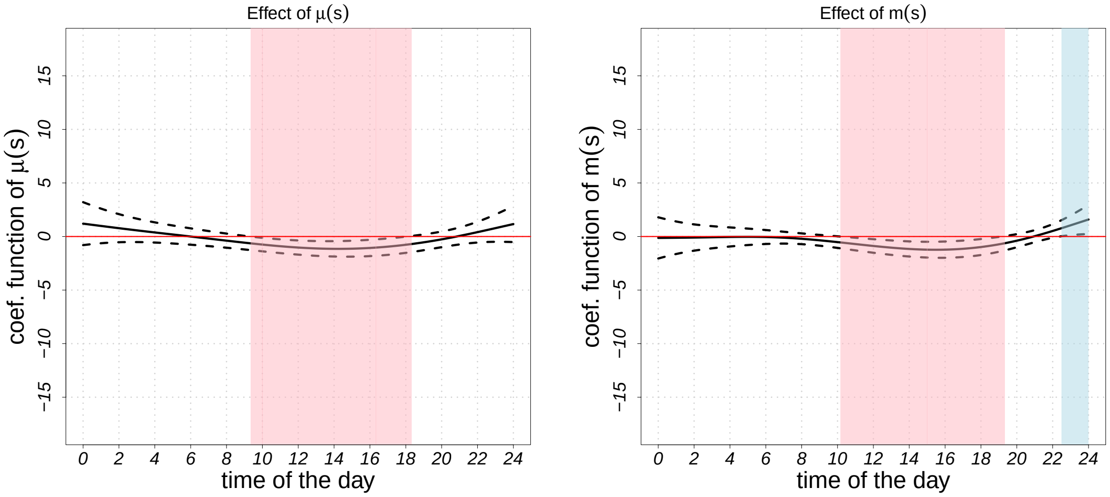


Figure S.5: The estimated regression coefficients for models employing temporally local summary functions such as mean, and mode based on activity counts (in logarithmic scale) are derived through scalar-on-function regressions to analyze EDSS scores. These functions are generated using log-activity counts during periods of activity. The solid black curve represents the estimated coefficient function, while dotted black lines denote the 95% confidence interval.

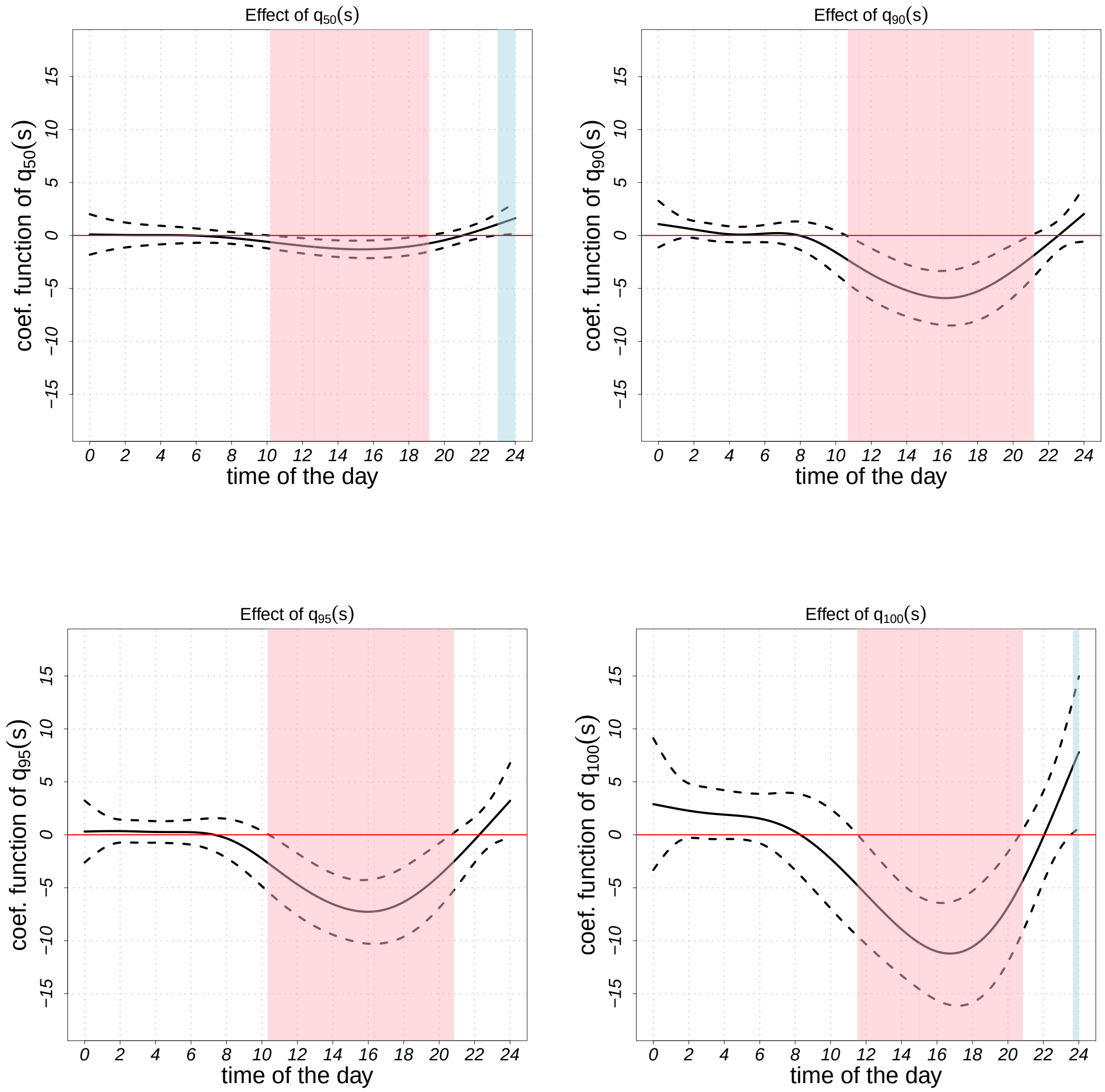


Figure S.6: The estimated regression coefficients for models employing temporally local quantile functions based on activity counts (in logarithmic scale) that are derived through scalar-on-function regressions to analyze EDSS scores. These functions are generated using log-activity counts during periods of activity. The solid black curve represents the estimated coefficient function, while dotted black lines denote the 95% confidence interval.

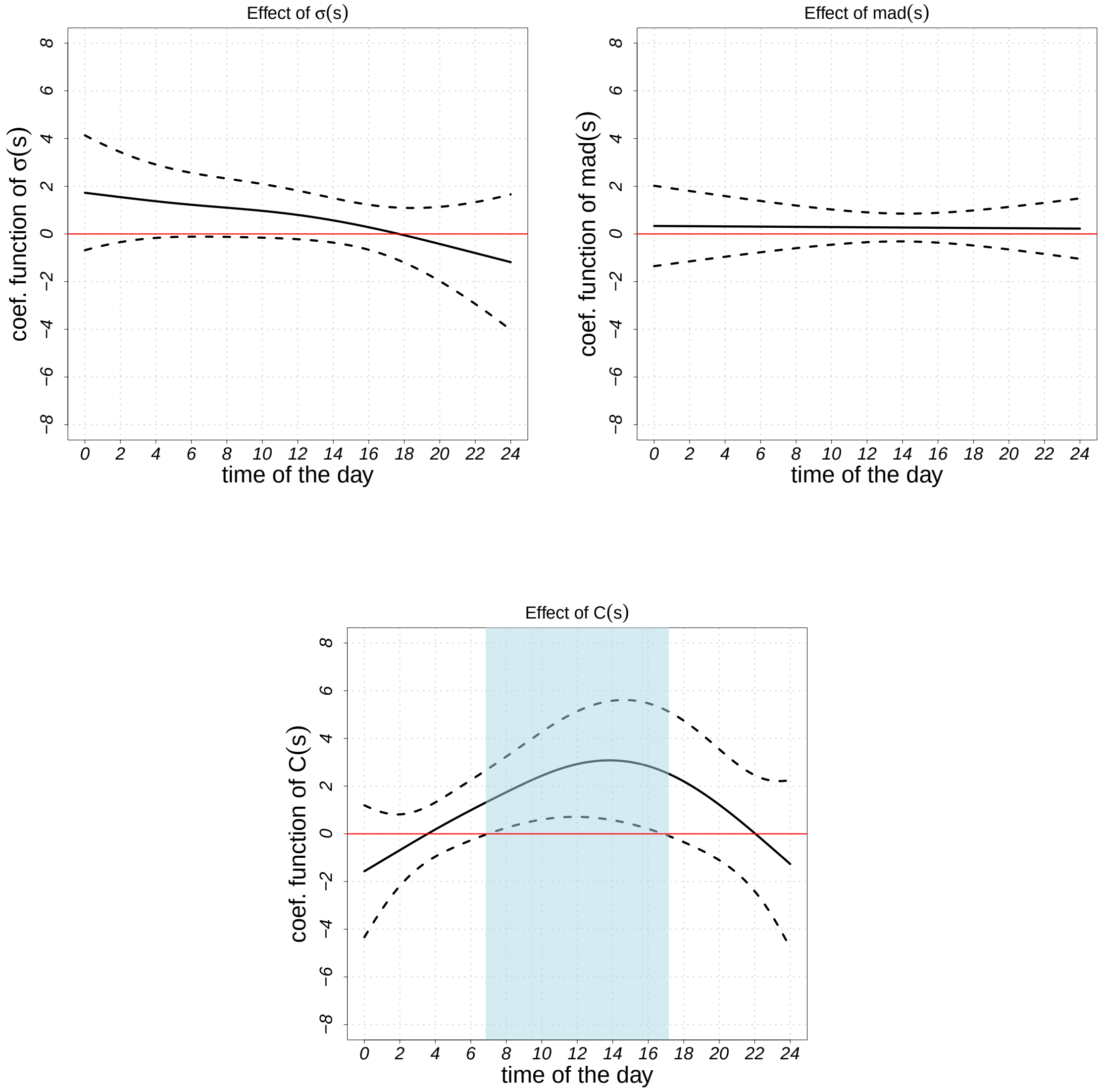


Figure S.7: The estimated regression coefficients for models employing temporally local variability functions such as standard deviation, median absolute deviation, and coefficient of variation based on activity counts (in logarithmic scale) are derived through scalar-on-function regressions to analyze EDSS scores. These functions are generated using log-activity counts during periods of activity. The solid black curve represents the estimated coefficient function, while dotted black lines denote the 95% confidence interval.

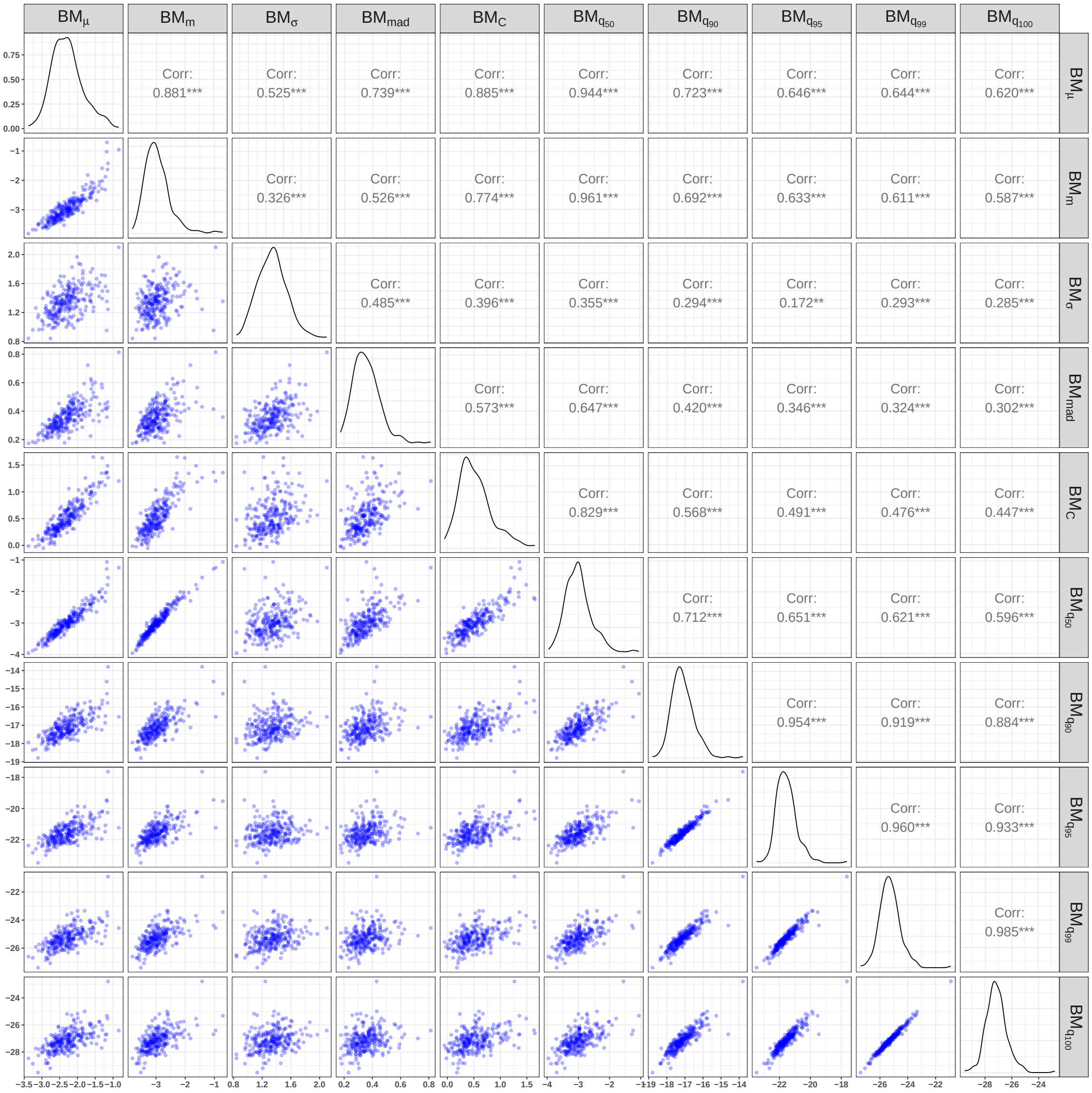


Figure S.8: Scatterplots of the estimated weighted scores corresponding to the predictor of different local distributional characteristic metrics, respectively. These local distributional characteristic metrics were calculated based on the physical activity counts in the logarithmic scale. All correlation coefficients indicate Spearman's rank correlation.

Table S.1: The results of modeling EDSS scores and different local distributional characteristic metrics of physical activity (in original scale) using SOFR with an adjustment for age, gender, and BMI. The standard deviation of the estimated coefficients for the scalar predictors is indicated in the parentheses.

| Choice of covariates | Intercept | Age | Sex (Male) | BMI | |
|---|---|---|---|---|---|
| mean, $\mu(s)$ | 3.374 *** | 0.029 * | -0.032 | -0.008 | $\widehat{\beta}_{\mu}(s)$*** |
| | ( 0.944) | (0.011) | (0.212) | (0.017) | |
| mode, $m(s)$ | 1.560$^{\cdot}$ | 0.037 ** | 0.069 | 0.001 | $\widehat{\beta}_{m}(s)^{\cdot}$ |
| | (0.896) | (0.012) | (0.220) | (0.017) | |
| standard deviation, $\sigma(s)$ | 5.178 *** | 0.023 * | -0.074 | -0.017 | $\widehat{\beta}_{\sigma}(s)$ *** |
| | (0.956) | (0.011) | (0.201) | (0.016) | |
| median abs. dev., $mad(s)$ | 2.646 ** | 0.033 ** | -0.060 | -0.003 | $\widehat{\beta}_{mad}(s)$ *** |
| | (0.914) | (0.012) | (0.216) | (0.017) | |
| coef. of variation, $\mathcal{C}(s)$ | 0.900 ** | 0.036 | 0.091 | 0.003 | $\widehat{\beta}_{\mathcal{C}}(s)^{\cdot}$ |
| | (1.076) | (0.012) | (0.221) | (0.017) | |
| 50th quantile, $q_{50}(s)$ | 2.088 * | 0.035 ** | 0.010 | -0.002 | $\widehat{\beta}_{q_{50}}(s)$* |
| | (0.910) | (0.012) | (0.219) | (0.017) | |
| 90th quantile, $q_{90}(s)$ | 4.631 *** | 0.026 * | -0.075 | -0.014 | $\widehat{\beta}_{q_{90}}(s)$*** |
| | (0.939) | (0.011) | (0.203) | (0.016) | |
| 95th quantile, $q_{95}(s)$ | 5.065 *** | 0.023 * | -0.078 | -0.015 | $\widehat{\beta}_{q_{95}}(s)$*** |
| | (0.958) | (0.011) | (0.202) | (0.016) | |
| 99th quantile, $q_{99}(s)$ | 5.456 *** | 0.019$^{\cdot}$ | -0.077 | -0.017 | $\widehat{\beta}_{q_{99}}(s)$*** |
| | (0.982) | (0.011) | (0.202) | (0.016) | |
| maximum, $q_{100}(s)$ | 5.613 *** | 0.018 | -0.068 | -0.017 | $\widehat{\beta}_{q_{100}}(s)$*** |
| | (0.992) | (0.011) | (0.202) | (0.016) | |

Signif. codes: '***' 0.001 '**' 0.01 '*' 0.05 '.' 0.1.

Table S.2: The results of modeling EDSS scores and different local distributional characteristic metrics of physical activity (in logarithmic scale) using SOFR with an adjustment for age, gender, and BMI. The standard deviation of the estimated coefficients for the scalar predictors is indicated in the parentheses.

| Choice of covariates | Intercept | Age | Sex (Male) | BMI | |
|---|---|---|---|---|---|
| mean, $\mu(s)$ | 4.180 *** | 0.030 * | 0.050 | -0.005 | $\widehat{\beta}_{\mu}(s)$*** |
| | (1.169) | (0.012) | (0.216) | (0.017) | |
| mode, $m(s)$ | 4.516 *** | 0.034 ** | 0.066 | -0.001 | $\widehat{\beta}_{m}(s)$*** |
| | (1.134) | (0.012) | (0.214) | (0.017) | |
| standard deviation, $\sigma(s)$ | 0.007 | 0.037 ** | 0.099 | 0.001 | $\widehat{\beta}_{\sigma}(s)$ |
| | (1.151) | (0.012) | (0.220) | (0.017) | |
| median abs. dev., $mad(s)$ | 0.783 | 0.039 ** | 0.072 | 0.004 | $\widehat{\beta}_{mad}(s)$ |
| | (0.896) | (0.012) | (0.223) | (0.017) | |
| coef. of variation, $\mathcal{C}(s)$ | 0.930 | 0.033 ** | 0.114 | 0.003 | $\widehat{\beta}_{\mathcal{C}}(s)$* |
| | (1.027) | (0.012) | (0.221) | (0.017) | |
| 50th quantile, $q_{50}(s)$ | 4.669 *** | 0.032 ** | 0.047 | -0.002 | $\widehat{\beta}_{q_{50}}(s)$*** |
| | (1.188) | (0.012) | (0.215) | (0.017) | |
| 90th quantile, $q_{90}(s)$ | 19.316 *** | 0.027 * | -0.108 | -0.008 | $\widehat{\beta}_{q_{90}}(s)$*** |
| | (2.961) | (0.011) | (0.208) | (0.016) | |
| 95th quantile, $q_{95}(s)$ | 23.787 *** | 0.026 * | -0.095 | -0.007 | $\widehat{\beta}_{q_{95}}(s)$*** |
| | (3.364) | (0.011) | (0.205) | (0.016) | |
| 99th quantile, $q_{99}(s)$ | 27.931 *** | 0.021· | -0.083 | -0.012 | $\widehat{\beta}_{q_{99}}(s)$*** |
| | (3.652) | (0.011) | (0.199) | (0.015) | |
| maximum, $q_{100}(s)$ | 29.981 *** | 0.018· | -0.084 | -0.013 | $\widehat{\beta}_{q_{100}}(s)$*** |
| | (3.796) | (0.011) | (0.197) | (0.015) | |

Signif. codes: '***' 0.001 '**' 0.01 '*' 0.05 '.' 0.1.
